# Influenza Antibody Levels Associated with Laboratory-Confirmed Influenza in a Test-Negative Study Design, US Flu VE Network, November 2018–May 2019

**DOI:** 10.64898/2026.03.24.26349239

**Authors:** Brendan Flannery, Jessie R. Chung, Crystal Holiday, Stacie Jefferson, Manjusha Gaglani, Kempapura Murthy, Richard K. Zimmerman, Mary Patricia Nowalk, Michael L. Jackson, Karen J. Wernli, Arnold S. Monto, Emily T. Martin, Huong Q. Nguyen, Joshua G. Petrie, Emma K. Noble, Kelsey M. Sumner, Lauren Grant, Zhu-Nan Li, Min Z. Levine

**Affiliations:** Centers for Disease Control and Prevention, Atlanta, GA, USA; Baylor Scott & White Health, Temple, TX, USA; Baylor College of Medicine, Temple, TX, USA; University of Pittsburgh, Pittsburgh, PA, USA; Kaiser Permanente Washington Health Research Institute, Seattle, WA, USA; University of Michigan School of Public Health, Ann Arbor, MI, USA; Marshfield Clinic Research Institute, Marshfield, WI, USA

**Author notes:** CORRESPONDING AUTHOR: Brendan Flannery, Address: U.S. Centers for Disease Control and Prevention, 1600 Clifton Rd Mailstop H24-7, Atlanta, GA 30329, USA.

**Keywords:** Influenza, Test-negative study, Influenza virus infection, Hemagglutinin, Neuraminidase

## Abstract

**Background:** We assessed associations between antibody concentrations within 7 days of symptom onset and testing positive for influenza virus infection among outpatients enrolled in a test-negative study.

**Methods:** From November 2018─May 2019, study sites in five states obtained serum and respiratory specimens from outpatients aged ≥18 years presenting with acute respiratory illness. Respiratory specimens were tested for influenza virus, and viral clades were identified by genomic sequencing. We measured influenza antibody titers against vaccine and circulating viruses by hemagglutination inhibition (HI), microneutralization (MN) and neuraminidase inhibition (NAI) assays. Percent of patients with HI, MN and NAI titers ≥10 and ≥40 were compared among patients with and without influenza-associated illness, and reduction in odds of confirmed influenza at increasing HI, MN and NAI antibody titers was estimated using logistic regression adjusting for influenza vaccination status and time since beginning of influenza season.

**Results:** Among 175 patients with confirmed influenza virus infection, including 112 with influenza A(H1N1)pdm09 and 63 with A(H3N2) (44 clade 3C.3a), and 130 test-negative control patients, higher antibody titers against influenza hemagglutinin or neuraminidase proteins at enrollment were associated with lower odds of influenza virus infection. HI and MN antibody titers against circulating viruses were more strongly associated with protection than titers against vaccine reference viruses. Odds of A(H1N1)pdm09 infection were 44% and 54% lower for each two-fold increase in A(H1N1)pdm09 HI or NAI titer, respectively. Odds of A(H3N2) infection were 46% and 30% lower, respectively, for each two-fold increase in MN or NAI titer against circulating A(H3N2) virus clade. NAI titers were independently associated with lower odds of influenza A(H1N1)pdm09 and A(H3N2) after controlling for HI titer.

**Conclusion:** Higher influenza antibody titers against circulating viruses were associated with lower likelihood of influenza virus infection among adult patients with acute respiratory illness.

**SUMMARY:** From November 2018─May 2019, we assessed the association between antibody concentrations during acute illness and laboratory-confirmed influenza among adult patients enrolled in a test-negative study in five US states. We found that higher influenza antibody titers were associated with lower likelihood of symptomatic influenza virus infection.

## INTRODUCTION

Effectiveness of seasonal influenza vaccine varies by season, circulating influenza viruses and population immunity [1]. During the 2018-2019 season, a test-negative study of influenza vaccine effectiveness reported protection against influenza A(H1N1)pdm09-associated illness, but no significant protection against medically-attended illness due to influenza A(H3N2) virus during the second half of the season when antigenically drifted A(H3N2) viruses predominated [2]. Influenza vaccine effectiveness (VE) and protection have generally been lower when circulating influenza viruses differ antigenically from vaccine reference viruses [3, 4]. Establishing correlations between immune responses to influenza vaccination and protection against medically attended influenza may contribute to improved vaccine strain selection.

Test-negative design (TND) studies are widely used to evaluate influenza vaccine effectiveness [5–7]. Measurement of specific antibody levels in serum specimens collected from patients presenting to healthcare with acute respiratory illness [8–11] serves as a proxy for pre-existing antibody levels at the time of exposure [8, 12]. In TND studies of influenza vaccine effectiveness, symptomatic patients who seek medical care for an acute respiratory illness are systematically enrolled and tested for influenza virus infection [6]. Odds of influenza virus infection are compared among patients vaccinated versus unvaccinated against influenza. Reduction in the odds of laboratory-confirmed influenza provides an estimate of VE against disease endpoints. Following convalescence from illness, immune response to infection is measured by comparing antibody titers during acute illness with convalescent-phase titers.

Genetic characterization of influenza viruses isolated from patients informs selection of serology antigens to measure immune responses specific to viral clade or genetic group [2, 13]. Odds of symptomatic infection may be compared at increasing titers of antibodies against the infecting virus [8, 10]. In this report, we assessed associations between symptomatic influenza virus infection, antibodies against vaccine and circulating influenza viruses during acute respiratory illness and antibody response to infection among patients enrolled in an influenza vaccine effectiveness study.

## MATERIALS AND METHODS

### Study population and sample collection

Outpatients aged ≥18 years presenting within 7 days of onset of respiratory illness were enrolled from participating healthcare facilities across five study sites in the US Influenza Vaccine Effectiveness Network, as previously described [2, 14]. Epidemiologic data collected from enrolled patients included patient age, presence of underlying medical conditions, date of illness onset, reported symptoms and receipt of 2018-2019 influenza vaccine. Respiratory specimens (combined nasal and throat swabs) were tested by influenza real-time reverse-transcription polymerase chain reaction (RT-PCR). Influenza A viruses were subtyped by RT-PCR and characterized by whole genome sequencing at the US Centers for Disease Control and Prevention (CDC) to identify viral clade [2]. Sequence data for characterized influenza viruses were uploaded to the Global Initiative on Sharing All Influenza Data (Supplementary Material). Viruses were classified into hemagglutinin clade based on phylogenetic analyses. Patients were classified as infected with influenza virus (cases) or uninfected (test-negative controls) based on RT-PCR result.

At enrollment, research staff at each study site collected blood specimens from participants who consented to a venous blood draw. Sera were separated and aliquoted at study sites and sent to CDC for serologic testing. Acute-phase blood specimens were defined as collected within 7 days of reported date of illness onset. Participants who tested RT-PCR positive for influenza virus infection at enrollment (case-patients) were scheduled for a second, convalescent-phase venous blood draw 21 or more days after reported symptom onset. Follow-up blood specimens were not obtained from test-negative control participants. This study was reviewed and approved by Institutional Review Boards at each of the five participating institutions.

### Hemagglutination inhibition assay (HI)

Sera were aliquoted and stored at −70DC and shipped on dry ice to CDC for laboratory analyses. HI assays were performed as described previously [15]. Serially 2-fold-diluted sera were tested in duplicate using 0.5% turkey red blood cells from an initial 1:10 dilution (lowest detectable HI titer). HI titer was defined as the reciprocal of the last dilution of serum that completely inhibited hemagglutination. HI assay viruses included 2018-2019 vaccine reference strain A/Michigan/45/2015(H1N1)pdm09 (clade 6B.1) grown in embryonated chicken eggs and A/Michigan/45/2015(H1N1)pdm09 cultured in Madin Darby canine kidney cells, or cell culture-propagated viruses belonging to the same genetic group of viruses isolated from patients. Test viruses were sequenced after propagation to verify absence of introduced genetic mutations.

### Microneutralization assay (MN)

Because A(H3N2) clade 3C.2a and 3C.3a viruses had reduced ability to hemagglutinate red blood cells, A(H3N2) antibody titers were determined using influenza virus microneutralization assays [16]. For MN assays, serial 2-fold dilutions from an initial 1:10 dilution (lowest detectable MN titer) were incubated with influenza viruses (100 50% tissue culture infective doses [TCID50]). The mixtures were infected with 1.5 x 10^4^ Madin-Darby canine kidney-SIAT1 (MDCK-SIAT1) cells per well, and viral infection was determined by ELISA. Neutralizing antibody titers were defined as the reciprocal of the highest dilution of serum samples that achieved at least 50% neutralization. MN assay viruses included 2018-2019 vaccine reference strain A/Singapore/2016(H3N2 clade 3C.2a.1) grown in embryonated chicken eggs or MDCK-SIAT1 cells and A/Kansas/2014(H3N2 clade 3C.3a) cultured in MDCK-SIAT1 representing the predominant circulating A(H3N2) virus.

### Neuraminidase inhibition assay (NAI)

Antibody concentrations that inhibited enzymatic activity of influenza viral neuraminidase were measured using enzyme-linked lectin assay (ELLA) with reassortant viruses containing neuraminidase from A/Michigan/45/2015(H1N1)pdm09 or A/Singapore/INFIMH-16-0019/2016(H3N2) with mismatched H6 hemagglutinin (i.e. rH6N1 or rH6N2), to avoid interference by HA-specific antibodies [17]. Patient serum was titrated in serial two-fold dilutions from an initial 1:10 dilution. NAI titers were calculated as the reciprocal of the highest serum dilution that inhibited neuraminidase activity by at least 50%. NAI titers were transformed and analyzed as described for HI and MN titers.

### Statistical analysis

To compare antibody levels during acute illness among influenza virus-infected case patients and uninfected test-negative control patients, we calculated geometric mean titers (GMT) with 95% confidence bounds and proportion of patients with HI and MN titers ≥10 and ≥40 against egg-propagated vaccine reference viruses or cell-propagated viruses belonging to the same viral clade as infecting strains, as well as proportions of NAI titers ≥10 and ≥40 against recombinant viruses. Proportions of HI, MN and NAI titers ≥10 and ≥40 were also compared by influenza vaccination status. We used chi-squared tests for statistical differences in proportions and t-tests for differences in GMT.

To quantify antibody response to infection, we calculated mean fold rise (MFR) with 95% confidence bounds as the ratio of serum antibody titers assayed in convalescent- versus acute-phase blood specimens, and proportion of seroconversion. For HI and MN assays, seroconversion was defined as ≥4-fold increase in antibody titers against the infecting viral clade antigen from acute- to convalescent-phase of illness, with a convalescent-phase titer ≥40. For NAI assays, fold increase was defined as the ratio of convalescent-phase to acute-phase antibody titers.

To compare odds of influenza virus infection among enrolled patients by antibody levels during the acute phase of illness, we calculated odds ratios of cases to test-negative control patients by HI, MN or NAI titer using logistic regression models adjusted for 2018-2019 influenza vaccination status and time since the beginning of influenza season, based on previous analyses [18, 19]. Percent change in relative odds of influenza virus infection was calculated as (1-adjusted odds ratio) x 100. Estimates were calculated separately for each assay and antigen.

All statistical analyses were performed using R version 4.4.0 (R Foundation for Statistical Computing, Vienna, Austria).

## RESULTS

From 23 November 2018 through 3 May 2019, we enrolled 6323 ambulatory patients aged ≥18 years 0-7 days after onset of respiratory symptoms. Serum specimens were collected from 305 patients for influenza A serology, including 112 patients infected with influenza A(H1N1)pdm09 (all 80 sequenced viruses were clade 6B.1A) and 63 with A(H3N2) clade 3C.3a (n=44 [92%] of 48 sequenced viruses) or 3C.2a/2a1 (n=4) viruses, and 130 influenza RT-PCR-negative patients (comparison group) (Figure 1).

**Figure 1.**
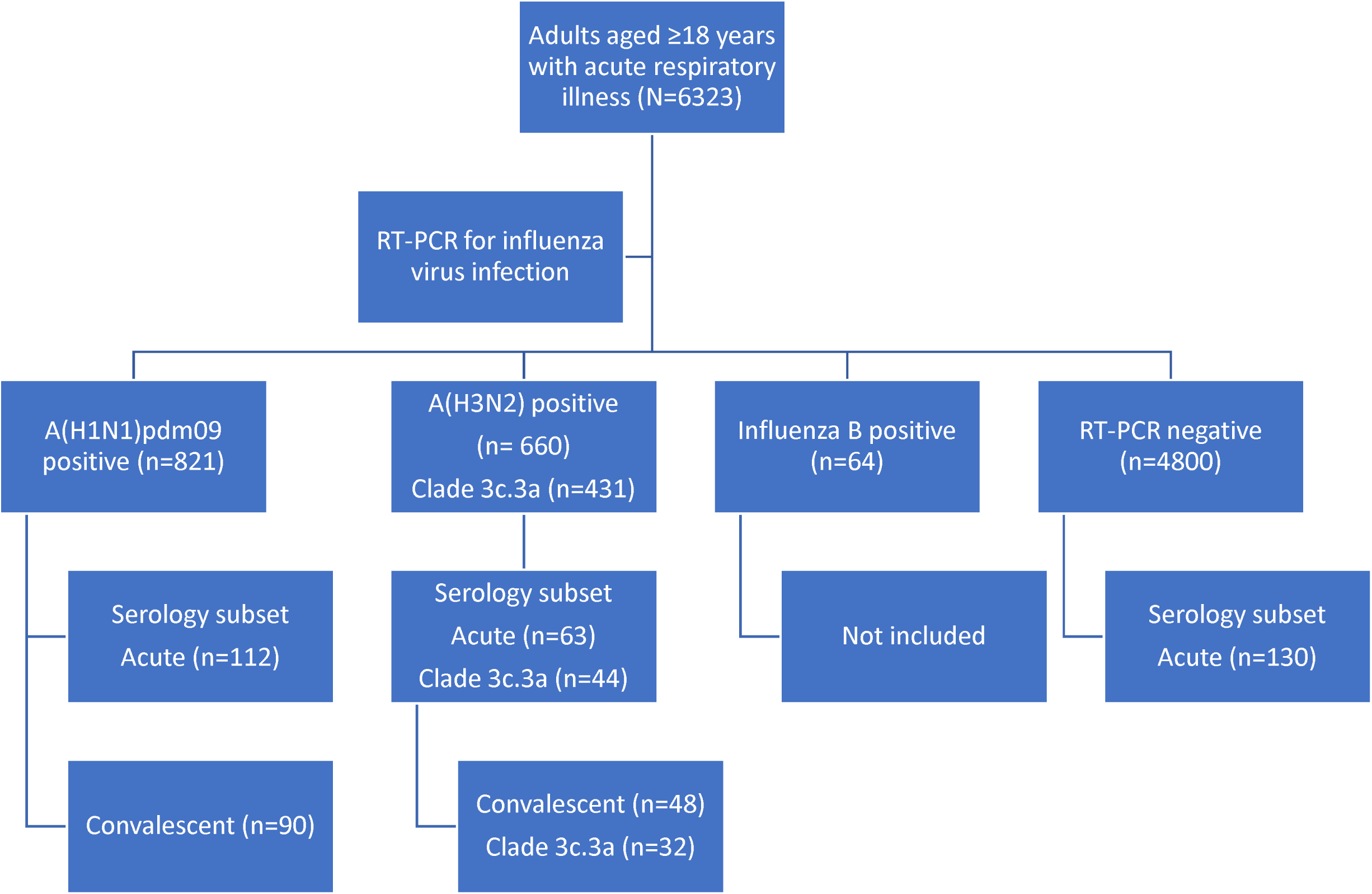
Study diagram of patients aged ≥18 years with acute respiratory illness enrolled in the serologic substudy of the test-negative design study of 2018-2019 influenza vaccine effectiveness. Numbers in parentheses are individual patients meeting criteria for inclusion in the serologic substudy by RT-PCR test result, virus clade and serum specimen collection.

Influenza B case patients were not included in the serologic substudy. Median age of patients was 47 years for A(H1N1)pdm09-positive cases and 51 years for both A(H3N2)-positive cases and test-negative controls (Table 1). In total, 68% of patients were female. Half of A(H1N1)pdm09-infected patients, two thirds of A(H3N2)-infected and 58% of non-influenza control patients had received 2018-2019 influenza vaccination. Acute-phase sera were collected a median of 4, 2, and 3 days (range, 0-7 days) after reported date of symptom onset for test-negative control patients, A(H1N1)pdm09-infected, and A(H3N2)-infected case patients, respectively. Convalescent-phase sera were collected after illness onset a median of 25 (range, 21 to 46) days from 90 influenza A(H1N1)pdm09-positive cases and 26 (range, 21 to 55) days from 48 influenza A(H3N2)-positive cases.

**Table 1.**
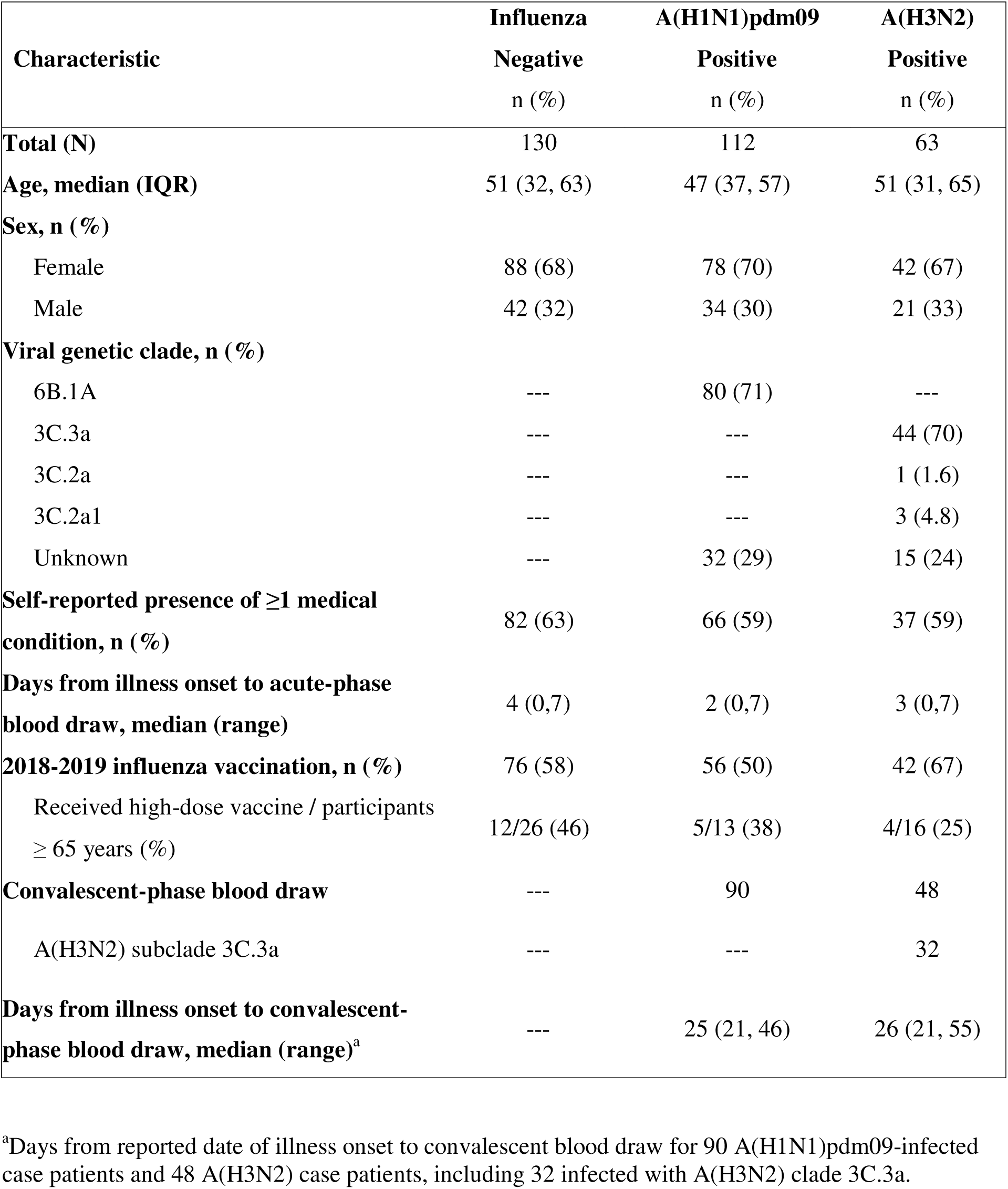
Characteristics of adult patients with RT-PCR confirmed influenza virus infection and RT-PCR test-negative control patients included in serologic substudy, 2018-2019.

### A(H1N1)pdm09

Geometric mean HI antibody titers at enrollment against egg-propagated A/Michigan/45/2015 vaccine reference virus and cell-propagated clade 6B.1A virus were lower among A(H1N1)pdm09 case patients than test-negative control patients (Table 2, Figure 2). Against the egg-propagated vaccine virus, 207 (86%) patients had detectable HI titers (≥10); 51 (46%) A(H1N1)pdm09-infected case patients compared with 92 (71%) uninfected test-negative patient controls had HI titers ≥40. Against cell-propagated clade 6B.1A virus, 199 (82%) patients had HI titers ≥10; 82 (73%) A(H1N1)pdm09 case patients compared with 117 (90%) control patients had HI titers ≥40. A(H1N1)pdm09 case patients also had lower geometric mean NAI antibody titers at enrollment against neuraminidase from A(H1N1)pdm09 compared with test-negative control patients. In all, 213 (88%) patients had acute-phase NAI titers ≥10; 44 (39%) A(H1N1)pdm09 case patients compared with 110 (85%) control patients had NAI titers ≥40 against rH6N1 virus. Geometric mean HI titers against egg- and cell-propagated A(H1N1)pdm09, as well as NAI GMTs against rH6N1 virus, were higher during acute-phase illness among vaccinated versus unvaccinated, uninfected control patients (Supplemental Table S1).

**Figure 2.**
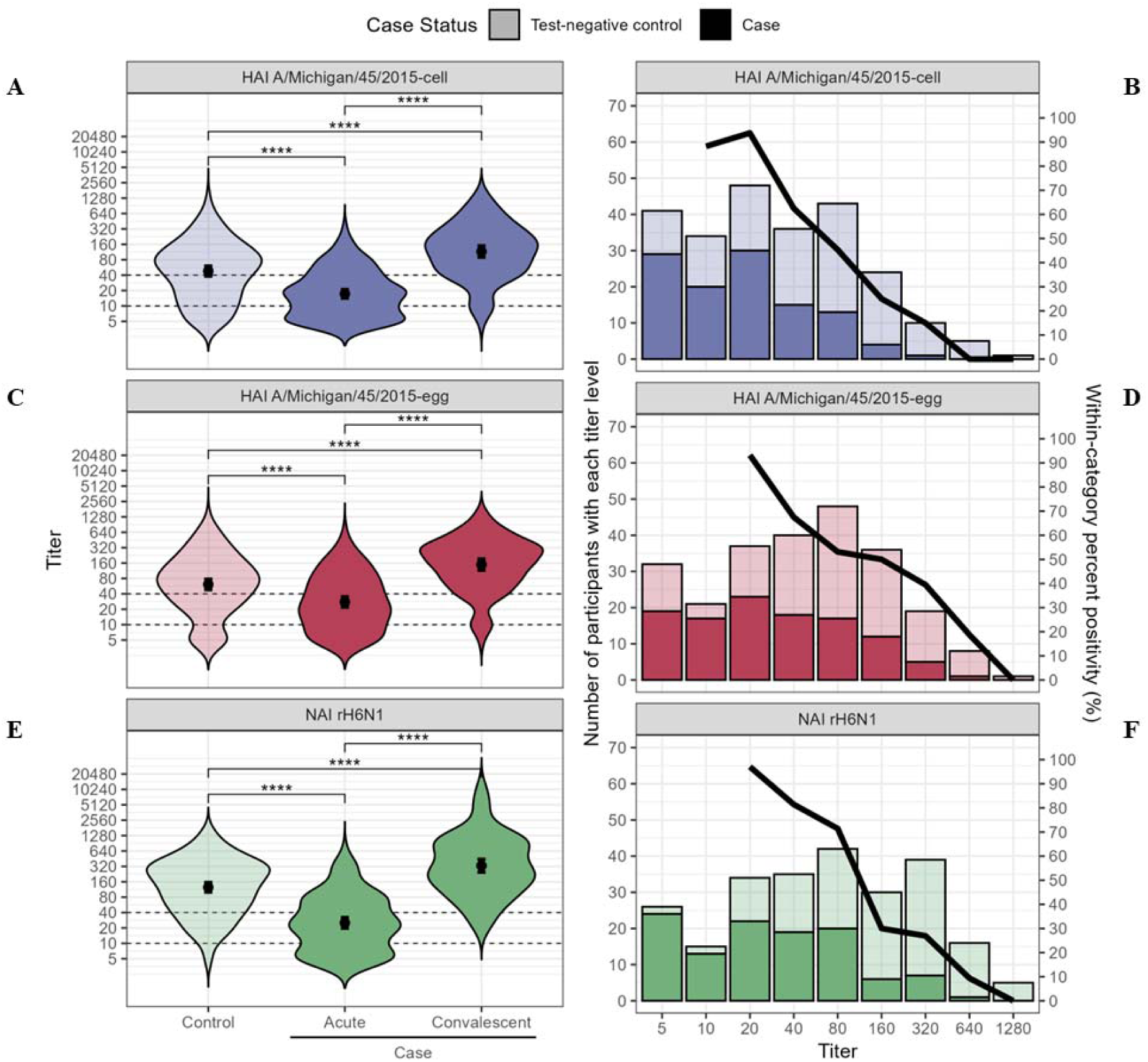
Association between influenza A(H1N1)pdm09 hemagglutination inhibition (HI) and neuraminidase inhibition (NAI) titers and influenza A(H1N1)pdm09 virus infection. **A-D.** Distribution of HI titers at time of outpatient enrollment (0-7 days after symptom onset) among uninfected test-negative control patients versus infected patients with RT-PCR confirmed A(H1N1)pdm09 clade 6B.1A virus infection, and convalescent-phase HI titers among case patients 24 to 42 days after illness onset. **A.** Violin plot of HI titer against cell culture-propagated A/Michigan/45/2015 virus (antigenically similar to circulating A(H1N1)pdm09 clade 6B.1A), and **B.** Percent A(H1N1)pdm09 positivity by HI titer against circulating viral clade. **C.** Violin plot of HI titer against egg-propagated A/Michigan/45/2015 vaccine reference virus, and **D.** Percent A(H1N1)pdm09 positivity by HI titer against egg-propagated vaccine reference virus. **E.** Distribution of acute-phase NAI titer by enzyme linked lectin assay (ELLA) with recombinant A(H6N1) virus containing A/California/9/2009 neuraminidase antigen comparing uninfected test-negative control patients, infected case patients during acute illness and convalescent-phase sera. **F.** Percent A(H1N1)pdm09 positivity by NAI titer. Bars indicate geometric mean titer and 95% confidence interval. Dashed lines indicate titers of 10 and 40. Asterisk indicates p-value for t-test of difference in geometric mean antibody titers. ****: p<0.001.

**Table 2.**
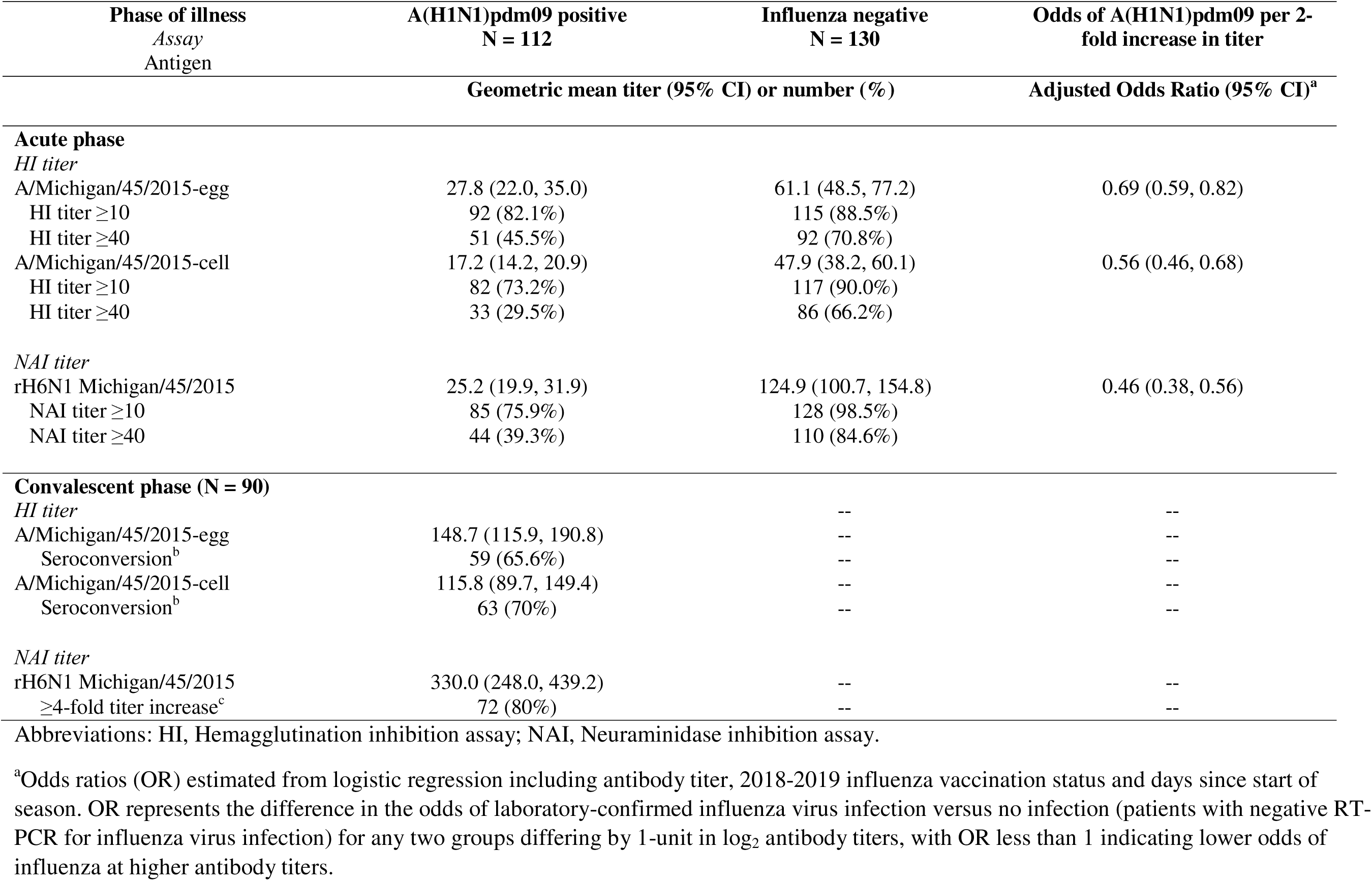
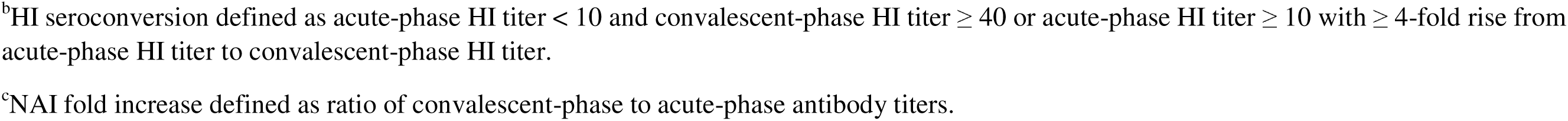
Association between hemagglutination inhibition and neuraminidase inhibition assay antibody titers during acute illness and influenza A(H1N1)pdm09 virus infection among patients with acute respiratory illness and antibody seroconversion among A(H1N1)pdm09-infected patients.

| Phase of illness<br>*Assay*<br>Antigen | A(H1N1)pdm09 positive<br>N = 112 | Influenza negative<br>N = 130 | Odds of A(H1N1)pdm09 per 2-fold increase in titer |
| --- | --- | --- | --- |
|  | Geometric mean titer (95% CI) or number (%) |  | Adjusted Odds Ratio (95% CI)<sup>a</sup> |
| **Acute phase** |  |  |  |
| *HI titer* |  |  |  |
| A/Michigan/45/2015-egg | 27.8 (22.0, 35.0) | 61.1 (48.5, 77.2) | 0.69 (0.59, 0.82) |
| HI titer ≥10 | 92 (82.1%) | 115 (88.5%) |  |
| HI titer ≥40 | 51 (45.5%) | 92 (70.8%) |  |
| A/Michigan/45/2015-cell | 17.2 (14.2, 20.9) | 47.9 (38.2, 60.1) | 0.56 (0.46, 0.68) |
| HI titer ≥10 | 82 (73.2%) | 117 (90.0%) |  |
| HI titer ≥40 | 33 (29.5%) | 86 (66.2%) |  |
| *NAI titer* |  |  |  |
| rH6N1 Michigan/45/2015 | 25.2 (19.9, 31.9) | 124.9 (100.7, 154.8) | 0.46 (0.38, 0.56) |
| NAI titer ≥10 | 85 (75.9%) | 128 (98.5%) |  |
| NAI titer ≥40 | 44 (39.3%) | 110 (84.6%) |  |
| **Convalescent phase (N = 90)** |  |  |  |
| *HI titer* |  | -- | -- |
| A/Michigan/45/2015-egg | 148.7 (115.9, 190.8) | -- | -- |
| Seroconversion<sup>b</sup> | 59 (65.6%) | -- | -- |
| A/Michigan/45/2015-cell | 115.8 (89.7, 149.4) | -- | -- |
| Seroconversion<sup>b</sup> | 63 (70%) | -- | -- |
| *NAI titer* |  |  |  |
| rH6N1 Michigan/45/2015 | 330.0 (248.0, 439.2) | -- | -- |
| ≥4-fold titer increase<sup>c</sup> | 72 (80%) | -- | -- |

Abbreviations: HI, Hemagglutination inhibition assay; NAI, Neuraminidase inhibition assay.
<sup>a</sup>Odds ratios (OR) estimated from logistic regression including antibody titer, 2018-2019 influenza vaccination status and days since start of season. OR represents the difference in the odds of laboratory-confirmed influenza virus infection versus no infection (patients with negative RT-PCR for influenza virus infection) for any two groups differing by 1-unit in log<sub>2</sub> antibody titers, with OR less than 1 indicating lower odds of influenza at higher antibody titers.
<sup>b</sup>HI seroconversion defined as acute-phase HI titer < 10 and convalescent-phase HI titer ≥ 40 or acute-phase HI titer ≥ 10 with ≥ 4-fold rise from acute-phase HI titer to convalescent-phase HI titer.
<sup>c</sup>NAI fold increase defined as ratio of convalescent-phase to acute-phase antibody titers.

From enrollment to convalescent follow-up among A(H1N1)pdm09-infected case patients, geometric mean HI titers against cell-propagated A/Michigan/45/2015 increased from 17.2 (CI: 14.2–20.9) to 115.8 (CI: 89.7–149.4) (Table 2); 63 (70%) A(H1N1)pdm09 case patients with paired acute- and convalescent-phase sera showed seroconversion by HI assay against cell-propagated clade 6B.1A virus, and 72 (80%) showed ≥4-fold increase in rH6N1 NAI titer. Geometric mean NAI titers against rH6N1 increased 13-fold from 25.2 (CI: 19.9–31.9) to 330.0 (CI: 248.0–439.2) (Table 2). Among 20 case patients with acute-phase HI titers <10, 19 (73%) showed seroconversion with convalescent tiers ≥40. For those with acute-phase NAI titers <10, 18 (86%) had a four-fold or greater NAI titer rise.

From logistic regression models controlling for 2018-2019 vaccination status and time since the beginning of the influenza season, each two-fold increase in acute-phase serum HI titer against the cell-propagated clade 6B.1A virus corresponded to a 44% reduction in odds for confirmed A(H1N1)pdm09 virus infection (OR, 0.56; 95% CI, 0.46-0.68) (Table 2). Each two-fold increase in NAI titer against rH6N1 virus corresponded to 54% lower odds for A(H1N1)pdm09 infection (OR, 0.46; 95% CI, 0.38-0.56). A(H1N1)pdm09-infected patients were more likely than uninfected patients to have acute-phase serum HI and NAI titers ≤40 (Figure 4A).

### A(H3N2)

At enrollment, patients infected with A(H3N2) clade 3C.3a viruses had lower geometric mean MN antibody titers than test-negative control patients against cell-propagated A/Kansas/14/2017 (clade 3C.3a) virus but not egg-propagated A/Singapore/INFIMH-16-0019/2016 (clade 3C.2a.1) vaccine reference virus (Table 3, Figure 3). Against the egg-propagated clade 3C.2a.1 vaccine virus, 164 (94%) patients had detectable MN titers (≥10); 30 (68%) A(H3N2) clade 3C.3a-infected case patients compared with 101 (78%) uninfected test-negative patient controls had MN titers ≥40 (Table 3). Against cell-propagated clade 3C.3a virus, 103 (59%) patients had MN titers ≥10; 6 (14%) A(H3N2) clade 3C.3a case patients compared with 55 (42%) control patients had MN titers ≥40. A(H3N2) clade 3C.3a-infected patients had similar geometric mean NAI antibody titers against rH6N2 virus compared with test-negative control patients. In all, 171 (98%) patients had acute-phase NAI titers ≥10; 38 (86%) A(H3N2) clade 3C.3a-infected case patients compared with 112 (86%) control patients had NAI titers ≥40 against rH6N2 virus. Geometric mean MN titers against egg- and cell-propagated A(H3N2), but not NAI GMTs against recombinant A(H6N2) virus, were higher during acute illness among vaccinated versus unvaccinated negative control patients (Supplemental Table S1).

**Figure 3.**
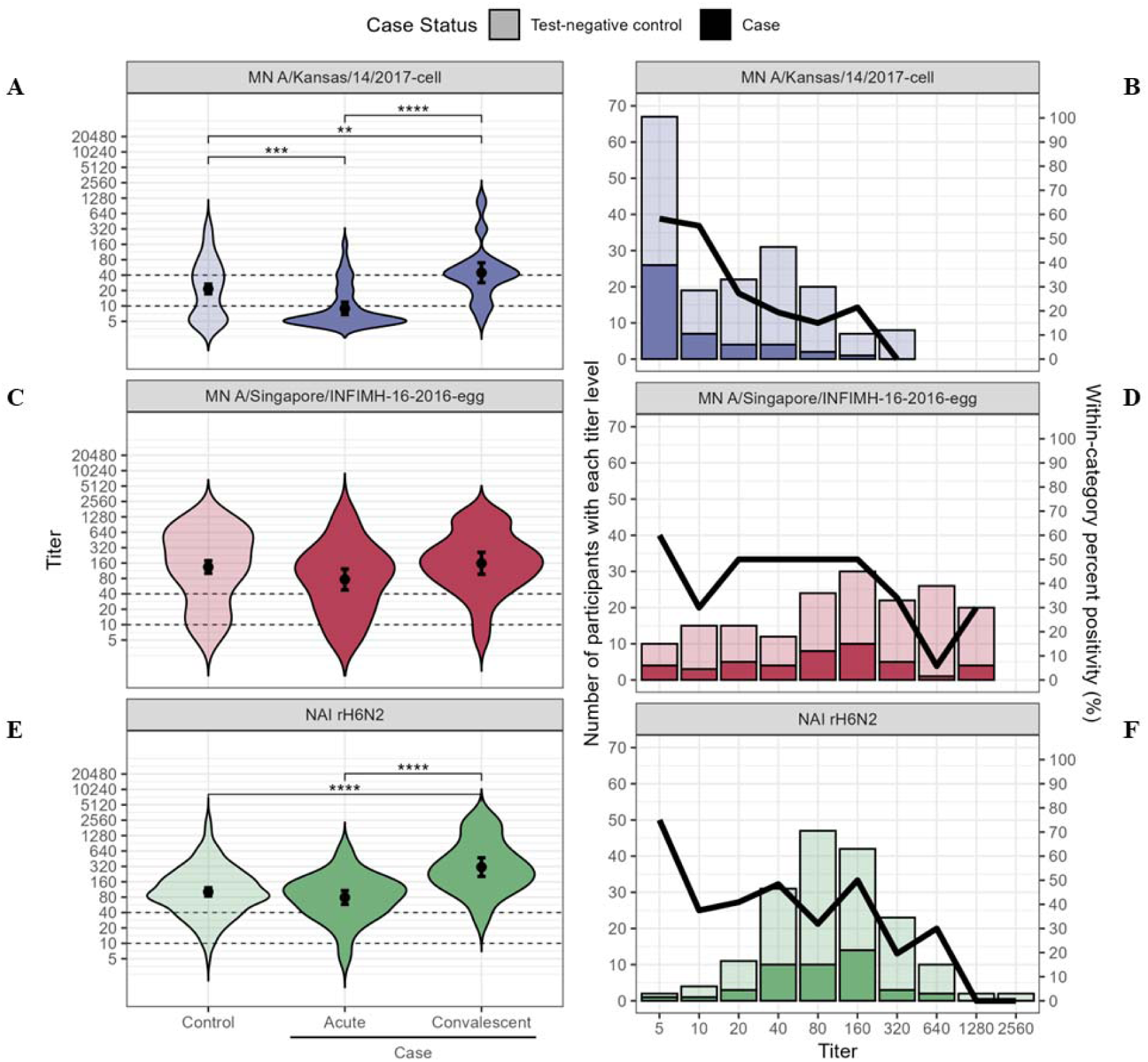
Association between influenza A(H3N2) microneutralization (MN) and neuraminidase inhibition (NAI) titers and influenza A(H3N2) clade 3C.3a-virus infection. A-D. Distribution of MN titers at time of outpatient enrollment (0-7 days after symptom onset) among uninfected test-negative control patients versus infected patients with RT-PCR confirmed A(H3N2) clade 3C.3a virus infection, and convalescent-phase MN titers among case patients 24 to 42 days after illness onset. **A.** Violin plot of MN titer against cell culture-propagated A/Kansas/14/2017 virus (antigenically similar to circulating A(H3N2) clade 3C.3a), and **B.** Percent A(H3N2) positivity by MN titer against circulating viral clade. **C.** Violin plot of MN titer against egg-propagated A/Singapore/INFIMH-16-0019/2016 vaccine reference virus, and **D.** Percent A(H3N2) positivity by MN titer against egg-propagated vaccine reference virus. **E.** Distribution of acute-phase NAI titer by enzyme linked lectin assay (ELLA) with recombinant A(H6N2) virus containing A/Singapore/INFIMH-16-0019/2016 neuraminidase antigen comparing uninfected test-negative control patients, infected case patients during acute illness and convalescent-phase sera. **F.** Percent A(H3N2) positivity by NAI titer. Bars indicate geometric mean titer. Bars indicate geometric mean titer and 95% confidence interval. Dashed lines indicate titers of 10 and 40. Asterisk indicates p-value for t-test of difference in geometric mean antibody titers. **: p<0.01; ****: p<0.005; ****: p<0.001.

**Figure 4.**
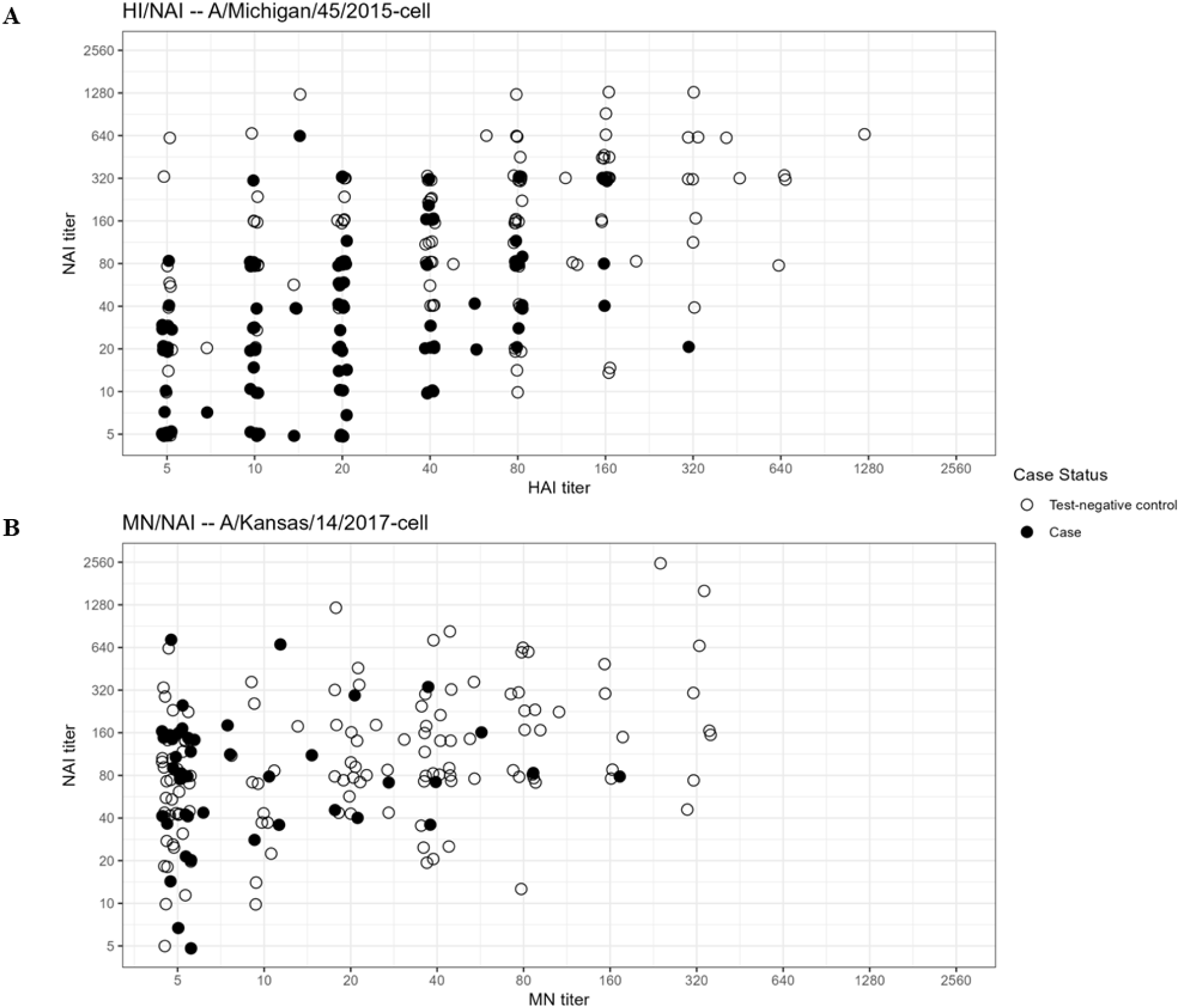
Distribution of hemagglutination inhibition (HI) or microneutralization (MN) and neuraminidase inhibition (NAI) assay titers among case patients with laboratory confirmed influenza virus infection or uninfected control patients with non-influenza respiratory illness. (**A**) HI titer against cell culture-propagated A/Michigan/45/2015 virus (antigenically similar to circulating A(H1N1)pdm09 clade 6B.1A) and NAI titer against recombinant A(H6N1) virus among A(H1N1)pdm09-infected patients (solid circles) or uninfected control patients (open circles); (**B**) MN titer against cell culture-propagated A/Kansas/14/2017 virus (antigenically similar to circulating A(H3N2) clade 3C.3a) and NAI titer against recombinant A(H6N2) virus among A(H3N2) clade 3C.3a-infected patients (solid circles) and test-negative control patients (open circles).

**Table 3.**
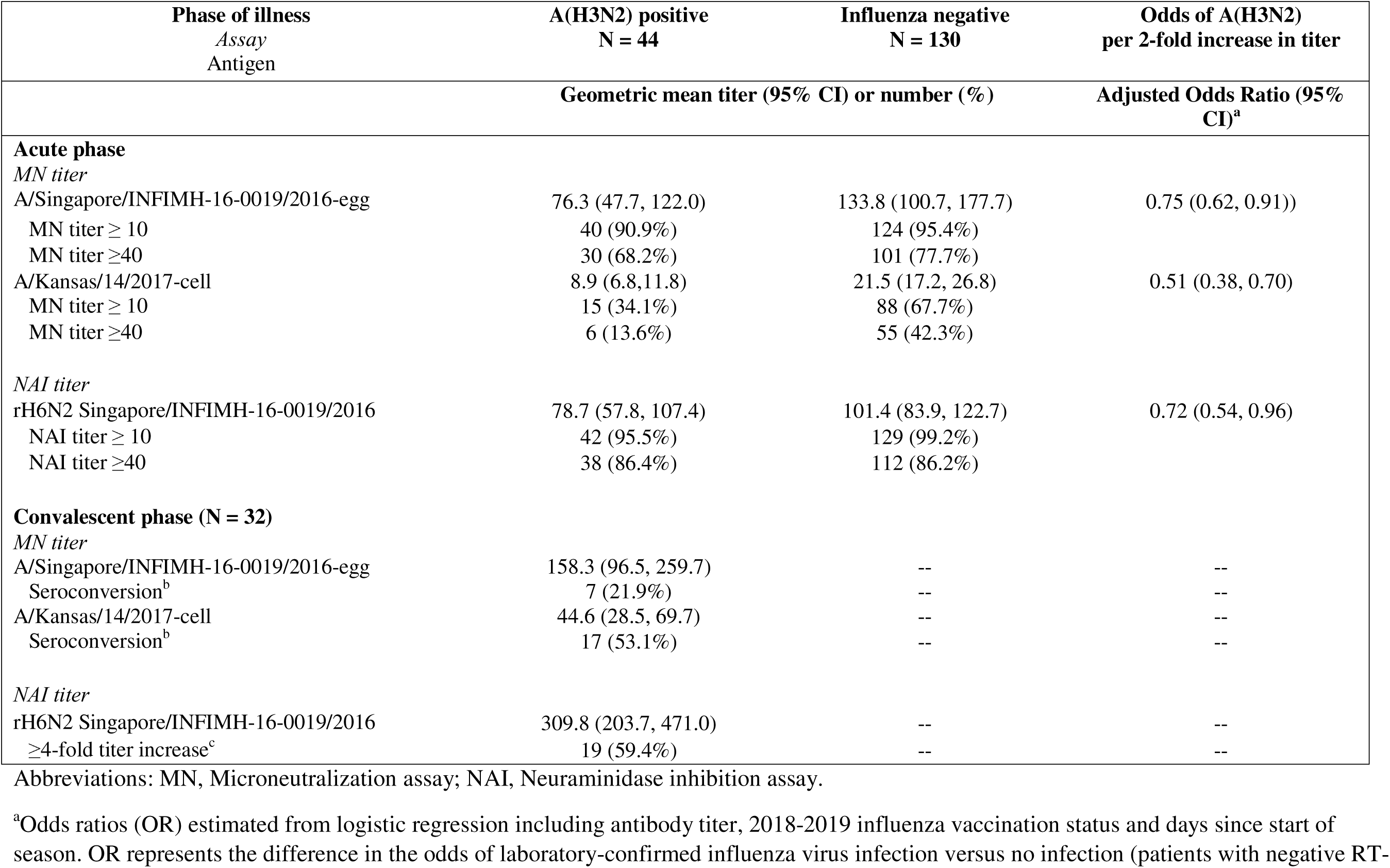
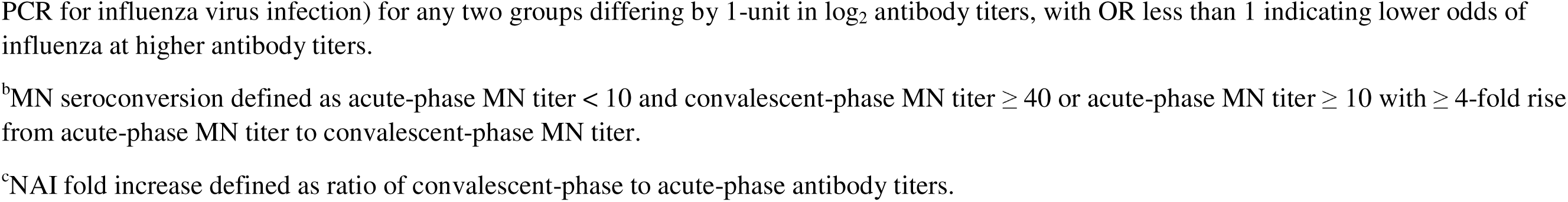
Association between influenza virus microneutralization and neuraminidase inhibition assay antibody titers during acute illness and influenza A(H3N2) virus infection among patients with acute respiratory illness and antibody seroconversion among A(H3N2) clade 3C.3a virus-infected patients.

| Phase of illness<br>*Assay*<br>*Antigen* | A(H3N2) positive<br>N = 44 | Influenza negative<br>N = 130 | Odds of A(H3N2)<br>per 2-fold increase in titer |
| --- | --- | --- | --- |
|  | Geometric mean titer (95% CI) or number (%) |  | Adjusted Odds Ratio (95% CI)<sup>a</sup> |
| **Acute phase** |  |  |  |
| *MN titer* |  |  |  |
| A/Singapore/INFIMH-16-0019/2016-egg | 76.3 (47.7, 122.0) | 133.8 (100.7, 177.7) | 0.75 (0.62, 0.91)) |
| MN titer ≥ 10 | 40 (90.9%) | 124 (95.4%) |  |
| MN titer ≥40 | 30 (68.2%) | 101 (77.7%) |  |
| A/Kansas/14/2017-cell | 8.9 (6.8, 11.8) | 21.5 (17.2, 26.8) | 0.51 (0.38, 0.70) |
| MN titer ≥ 10 | 15 (34.1%) | 88 (67.7%) |  |
| MN titer ≥40 | 6 (13.6%) | 55 (42.3%) |  |
| *NAI titer* |  |  |  |
| rH6N2 Singapore/INFIMH-16-0019/2016 | 78.7 (57.8, 107.4) | 101.4 (83.9, 122.7) | 0.72 (0.54, 0.96) |
| NAI titer ≥ 10 | 42 (95.5%) | 129 (99.2%) |  |
| NAI titer ≥40 | 38 (86.4%) | 112 (86.2%) |  |
| **Convalescent phase (N = 32)** |  |  |  |
| *MN titer* |  |  |  |
| A/Singapore/INFIMH-16-0019/2016-egg | 158.3 (96.5, 259.7) | -- | -- |
| Seroconversion<sup>b</sup> | 7 (21.9%) | -- | -- |
| A/Kansas/14/2017-cell | 44.6 (28.5, 69.7) | -- | -- |
| Seroconversion<sup>b</sup> | 17 (53.1%) | -- | -- |
| *NAI titer* |  |  |  |
| rH6N2 Singapore/INFIMH-16-0019/2016 | 309.8 (203.7, 471.0) | -- | -- |
| ≥4-fold titer increase<sup>c</sup> | 19 (59.4%) | -- | -- |

Abbreviations: MN, Microneutralization assay; NAI, Neuraminidase inhibition assay.
<sup>a</sup>Odds ratios (OR) estimated from logistic regression including antibody titer, 2018-2019 influenza vaccination status and days since start of season. OR represents the difference in the odds of laboratory-confirmed influenza virus infection versus no infection (patients with negative RT-
PCR for influenza virus infection) for any two groups differing by 1-unit in $\log_2$ antibody titers, with OR less than 1 indicating lower odds of influenza at higher antibody titers.
<sup>b</sup>MN seroconversion defined as acute-phase MN titer $< 10$ and convalescent-phase MN titer $\geq 40$ or acute-phase MN titer $\geq 10$ with $\geq 4$ -fold rise from acute-phase MN titer to convalescent-phase MN titer.
<sup>c</sup>NAI fold increase defined as ratio of convalescent-phase to acute-phase antibody titers.

From enrollment to convalescent follow-up among A(H3N2) clade 3C.3a-infected case patients, geometric mean MN titers against cell-propagated A/Kansas/14/2017 clade 3C.3a virus increased from 8.9 (CI: 6.8–11.8) to 44.6 (CI: 28.5–69.7) (Table 3); 17 (53%) A(H3N2) case patients with paired acute-and convalescent-phase sera showed seroconversion by MN assay against cell-propagated clade 3C.3a virus, and 19 (59%) showed ≥4-fold increase in rH6N2 NAI titer. Among 29 case patients with acute-phase MN titers <10, 11 (52%) showed seroconversion with convalescent tiers ≥40. One of two A(H3N2)-infected patients with acute-phase NAI titers <10 had a four-fold rise in rH6N2 NAI titer. Geometric mean rH6N2 NAI titers increased approximately four-fold from 78.7 (CI: 57.8–107.4) to 309.8 (CI: 203.7–471.0) (Table 3).

From logistic regression models controlling for 2018-2019 influenza vaccination status and time since the beginning of the influenza season, each two-fold increase in acute-phase serum MN titer against the cell-propagated A(H3N2) clade 3C.3a virus corresponded to a 49% reduction in odds for A(H3N2) virus infection (OR, 0.51; 95% CI, 0.38-0.70) (Table 3). Each two-fold increase in rH6N2 NAI titer corresponded to 28% lower odds ratio for A(H3N2) infection (OR, 0.72; 95% CI, 0.54, 0.96). A(H3N2)-infected patients were more likely than uninfected patients to have acute-phase serum MN and NAI titers ≤40 (Figure 4A).

## DISCUSSION

In this observational study using a test-negative design to estimate antibody titers associated with protection against medically attended influenza in adults, higher virus neutralization, hemagglutination and neuraminidase inhibition antibody titers at the time of enrollment were associated with lower odds of RT-PCR confirmed medically attended outpatient influenza virus infection. The strongest associations were observed when influenza viral antigens in serologic assays were antigenically similar to circulating viruses belonging to the same viral clade with which the case patient was infected. Higher A(H1N1)pdm09 hemagglutination inhibition antibodies and A(H3N2) neutralizing antibodies were associated with lower odds of A(H1N1)pdm09 or A(H3N2) infection, respectively. Among A(H1N1)pdm09 test-negative control patients, associations between influenza vaccination status and antibodies against circulating strains were consistent with observed vaccine effectiveness against A(H1N1)pdm09 clade 6B.1A-associated influenza. In contrast, despite higher geometric mean MN titers against A(H3N2) vaccine and clade 3C.3a viruses among influenza vaccinated compared with unvaccinated test-negative patients, low geometric mean MN titers overall may explain why vaccination was not associated with protection against A(H3N2) clade 3C.3a-associated illness during the 2018-2019 influenza season [2, 14].

Using influenza test-negative patients as a comparison group for influenza virus-infected case patients, we found that higher antibody titers were associated with lower odds of medically attended influenza illness. Studies from influenza vaccine trials have correlated virus neutralization and hemagglutination inhibition antibodies with influenza attack rates, demonstrating that HI and MN assay titers are serologic correlates of protection against symptomatic influenza virus infection [20–25]. Neuraminidase inhibiting antibodies have also been shown to correlate with protection and may act synergistically with HI or neutralizing antibodies [26–29]. However, randomized trials and cohort studies have limitations when evaluating protective antibody levels against antigenically drifted viruses. The test-negative design provides efficient enrollment of patients with laboratory-confirmed influenza (depending on proportion of influenza infected patients among those meeting enrollment criteria) and an uninfected comparison group of patients presenting with acute respiratory symptoms, which produces estimates similar to the exposure-proximal case-cohort method of determining antibody thresholds or serologic correlates of protection [8, 30, 31]. In this analysis, acute-phase antibody titers assessed at enrollment are used as indicators of antibody levels at the time of exposure, assuming that non-case patients were exposed to influenza viruses circulating in the community during the same time period. Higher antibody levels against influenza proteins representing circulating viral clades were associated with a lower likelihood of medically attended influenza virus infection. These results suggest that test-negative studies may provide a means of assessing immune correlations of protection by estimating antibody levels close to the time of infection. This may be particularly important when circulating influenza viruses differ antigenically from vaccine reference viruses, which are often used as serologic reference antigens to estimate correlate of protection thresholds based on post-vaccination antibody titers in randomized trials or cohort studies.

These findings are subject to several limitations. The serologic substudy was designed to assess applicability of the test-negative study design to measure antibody levels associated with protection against laboratory-confirmed influenza among adult outpatients. Patients who consented to participate in the serologic substudy were not randomly selected and did not represent all adult patients enrolled in the vaccine effectiveness study. Second, the serologic substudy did not recruit pediatric patients or hospitalized adults with acute respiratory illness. Additional serologic and cell mediated immune markers could be investigated among children [25, 32], as well as biomarkers associated with protection against severe disease [22, 33]. Third, acute-phase antibody titers may reflect early immune response in some influenza virus-infected individuals, resulting in an overestimation of antibody titers among case patients; however, studies of anamnestic response to re-infection with SARS-CoV-2 suggest minimal rise in serum antibody levels during the first week after symptom onset [8, 34]. Finally, except for the subset of 138 paired acute and convalescent samples, serum antibody titers were assessed at one time point during acute illness from patients in the serologic substudy.

In conclusion, using a test-negative design, we found that higher influenza antibody titers against circulating viruses were associated with lower likelihood of influenza virus infection among adult patients with acute respiratory illness. These results suggest a role for observational studies in evaluating antibody levels associated with protection against influenza. Harmonization of serologic assays and regular updates of serology antigens that are antigenically similar to circulating influenza viruses can contribute to vaccine strain selection. Serologic correlates of protection against influenza could provide endpoints to compare relative vaccine immunogenicity against circulating viruses. With multiple licensed and recommended influenza vaccines, observational studies incorporating serum collection for analysis can complement immunogenicity studies in evaluation of relative influenza vaccine effectiveness.

## Notes

### Acknowledgments

*Baylor Scott and White Health, Texas A&M University College of Medicine:* Michael Smith, Chandni Raiyani, Wencong Chen, Lydia Clipper, Kelsey Bounds, Amanda Drake, Teresa Ponder, Mary Kylberg, Natalie Settele, Jeremy Ray, Jennifer Thomas, Jamie Walkowiak, Renee Day, Madhava Beeram, John Erwin, and Alejandro Arroliga; *University of Pittsburgh Schools of the Health Sciences and University of Pittsburgh Medical Center:* Rose Azrak, G. K. Balasubramani, Todd M. Bear, Duane Eisaman, Heather Eng, Andrew Fackler, Edward Garofolo, Robert Hickey, Philip Iozzi, Monika Johnson, Stephanie Kirk, Jason A. Lyons, Donald B. Middleton, Krissy K. Moehling, Jonathan M. Raviotta, Evelyn C. Reis, Bret Rosenblum, Sean Saul, Theresa Sax, Michael Susick, Joe Suyama, Leonard F. Urbanski, Alexandra Weissman, and John V. Williams; *Kaiser Permanente Washington Health Research Institute:* Zoe Kappelman, Erika Kiniry, Lawrence Madziwa, Matt Nguyen, Suzie Park, C. Hallie Phillips, and Stacie Wellwood; *University of Michigan and Henry Ford Health System:* Allen Achkar, Elizabeth Alleman, Trinh Anh Minh, Habeeb Al-Shohatee, Gabriela Augustinaitis, Sarah Bauer, Danielle Carroll, Caroline K. Cheng, Robert Deblander III, Michelle Groesbeck, Emileigh Johnson, Anne Kaniclides, Armanda Kimberly, Jenna Kiryakos, Marym Kuril, Lois E. Lamerato, Ryan E. Malosh, Maria Matta, E. J. McSpadden, Madeleine Mendelow, Niharika Rajesh, Bryan Richardson, Stephanie Robinson, Hannah Segaloff, Caleb Sokolowski, Rachael Swanson, and Rachel Truscon; *Marshfield Clinic Research Institute:* Elizabeth Armagost, Theresa Balinghasay, Tamara Braund, Deanna Cole, Carrie Curtis, Tom Dalcher, Alicia Easley, Terry Foss, Wayne Frome, Hannah Gourdoux, Gregg Greenwald, Sherri Guzinski, Kayla Hanson, Linda Heeren, Lynn Ivacic, Marie Janz, Tara Johnson, Julie Karl, Jennifer King, Tamara Kronenwetter Koepel, Diane Kohnhorst, Sarah Kopitzke, Erik Kronholm, Marcia Lichtenwald, Carrie Marcis, Karen McGreevey, Jennifer Meece, Nidhi Mehta, Vicki Moon, Madalyn Palmquist, Nan Pan, Rebecca Pilsner, DeeAnn Polacek, Martha Presson, Lauren Putnam, Carla Rottscheit, Crystal Sabatke, Jacklyn Salzwedel, Megan Sauer, Julian Savu, Ram Shrestha, Elisha Stefanski, Patrick Stockwell, and Sandy Strey; *Centers for Disease Control and Prevention (CDC):* Angie Foust, Wendy Sessions, LaShondra Berman, Juliana DaSilva, and Shoshona Le.

### Disclaimer

The findings and conclusions in this report are those of the authors and do not necessarily represent the views of the U.S. Centers for Disease Control and Prevention (CDC). Some authors are federal employees of the United States government, and this work was prepared as part of their official duties. Title 17 U.S.C. 105 provides that ‘copyright protection under this title is not available for any work of the United States Government.’

### Financial support

This project was supported by the CDC through cooperative agreements with University of Michigan (grant U01 IP001034), the University of Pittsburgh (grant U01 IP001035), Kaiser Permanent Washington Health Research Institute (grant U01 IP001037), Marshfield Clinic Research Institute (grant U01 IP001038), and Baylor Scott & White Health (grant U01 IP001039). University of Pittsburgh also received funding from the National Institutes of Health (grant UL1TR001857).

### Potential conflicts of interest

R. K. Z., M. P. N. and E. T. M. have received consulting fees or research funding from Merck. R. K. Z., M. L. J. and A. S. M. have received research or grant funding from Sanofi Pasteur. M.P.N. has received research funding from AstraZeneca. A. S. M. has received personal fees from Novartis and Protein Sciences. E. T. M. has received research funds from Roche Pharmaceuticals. H. Q. N. has received research funding from Seqirus. All other authors report no potential conflicts.

### Data availability

Data produced in the present study are available upon reasonable request to the corresponding author.

## Data Availability

All data produced in the present study are available upon reasonable request to the authors

---

**Supplemental Table 1.** Hemagglutination inhibition, microneutralization and neuraminidase inhibition assay antibody titers against antigens representing vaccine and circulating influenza viruses among test-negative control patients with acute respiratory illness by vaccination status.

| Assay<br>Antigen | Vaccinated test-negative control patients<br>N = 76 |  | Unvaccinated test-negative control patients<br>N = 54 |  |
| --- | --- | --- | --- | --- |
|  | Geometric Mean Titer<br>(95% CI) | Titer ≥40, n (%) | Geometric Mean Titer<br>(95% CI) | Titer ≥40, n (%) |
| ***HI titer*** |  |  |  |  |
| A/Michigan/45/2015-egg | 102.4 (79.7, 131.5) | 64 (84.2%) | 29.6 (20.6, 42.5) | 28 (51.9%) |
| A/Michigan/45/2015-cell | 74.3 (57.1, 96.7) | 61 (80.3%) | 25.8 (18.3, 36.5) | 25 (46.3%) |
| ***MN titer*** |  |  |  |  |
| A/Singapore/INFIMH-16-0019/2016-egg | 286.8 (215.8, 381.2) | 71 (93.4%) | 45.8 (30.2, 69.3) | 30 (55.6%) |
| A/Kansas/14/2017-cell | 33.3 (25.2, 43.8) | 43 (56.6%) | 11.6 (8.5, 15.9) | 12 (22.2%) |
| ***NAI titer*** |  |  |  |  |
| rH6N1 | 168.2 (129.0, 219.4) | 70 (92.1%) | 82.1 (58.7, 114.8) | 40 (74.1%) |
| rH6N2 | 122.3 (98.6, 151.7) | 71 (93.4%) | 78.0 (55.5, 109.5) | 41 (75.9%) |

## Notes

### Author Declarations

This study was reviewed and approved by Institutional Review Boards at Baylor Scott & White Health, University of Pittsburgh, Kaiser Permanente Washington Health Research Institute, University of Michigan School of Public Health, and Marshfield Clinic Research Institute.

